# Outcome definition and index validity in cross-national comparisons of diet-attributable metabolic burden

**DOI:** 10.64898/2026.07.25.26358918

**Authors:** Shangze Huang, Xinyan Wang

## Abstract

**Background:** In three national surveys, no cross-national ranking of diet-attributable metabolic burden was identified: the apparent ordering moved with the definition of the outcome and with the validity of the dietary index. Both were tested.

**Subjects/Methods:** We harmonised individual-participant data from national surveys in the USA (NHANES 2017–2018), Korea (KNHANES 2015–2021) and Mexico (ENSANUT 2018–2019); 56,365 adults in total, recovered Mexican blood-pressure data from the raw files, and defined metabolic syndrome (MetS) with a common five-component ATP III rule. We estimated survey-weighted sex-specific associations of the energy-adjusted Dietary Inflammatory Index (E-DII) with MetS, country-specific population-attributable fractions (PAFs) on a common counterfactual scale, and the E-DII’s correlation with log high-sensitivity C-reactive protein (hsCRP). A quantitative bias analysis examined whether validity differences could reverse the country ordering.

**Results:** The E-DII–MetS association was larger in women (USA OR 1.23, 95% CI 0.98–1.55; Korea 1.078, 1.028–1.132; Mexico 1.074, 0.992–1.163; interaction P=0.080) but disappeared without blood pressure in the outcome (pooled 1.065, 0.982–1.155; P=0.13). Female PAFs were 15.0%, 7.6% and 3.8%; no pairwise difference was distinguishable (all P≥0.23). E-DII validity was 0.137 in US women versus 0.040 in Korean women (P=2.0×10−4); the validity ratio was 0.29, correction reversed the observed ordering, and its interval straddled the reversal threshold.

**Conclusions:** Neither the direction nor the magnitude of cross-national differences in diet-attributable MetS burden was identified; the ordering was bounded by outcome definition and by index validity. Cross-national burden comparisons require local validation of both.

## Introduction

Metabolic syndrome (MetS) affects roughly one third of adults worldwide and its prevalence rises steeply in women after the menopausal transition [1,2]. Chronic low-grade inflammation links diet to MetS [3,4], and the Dietary Inflammatory Index (DII) is the most widely used summary of the inflammatory potential of diet [5,6]. Meta-analyses of the DII–MetS association have moved from a null pooled estimate in 2018 (RR 1.01) to 1.23 in 2021, 1.24 in 2024 and 1.33 in 2026, with substantial heterogeneity (I^2^≈74%) [7–9]. That trajectory is usually read as accumulating evidence for a modest association, but it also documents that the pooled estimate is unstable across study sets and outcome definitions.

For prevention policy the relevant quantity is not the odds ratio but the population-attributable fraction (PAF): the share of disease burden that would not occur if the exposure distribution shifted. Cross-national comparisons of diet-attributable burden are now routine [10,11], and they carry two implicit assumptions. The first is **outcome comparability**: that MetS is ascertained the same way in each population. The second is **index validity**: that the dietary index measures the same inflammatory construct in each dietary culture. Both assumptions are testable, and both are routinely left untested. Where they fail, a country-level PAF difference may reflect measurement and definition rather than biology, and the ordering of countries may not be identified from the data at all.

Rockhill et al. established that population-attributable fractions (PAFs) are not fixed properties of an exposure: they shift with exposure prevalence and with the confounding structure of the population [17]. That argument holds the instrument fixed and varies the population. Our claim is logically prior: before a country-specific PAF can be recomputed anywhere, the index must measure the same construct and the outcome must mean the same thing in each population. Both premises are testable, and here both fail. The E-DII–hsCRP correlation was 0.137 in US women and 0.040 in Korean women, a validity ratio of 0.294; at a Korean coefficient of 0.055 the observed ordering reverses, and correction moved the point estimate across. The outcome behaved the same way: removing blood pressure abolished the female association. Bakhshimoghaddam et al. pooled component-specific odds ratios across countries, which presupposes exactly this commensurability [8]. Our contribution is an identifiability criterion and a testable formalisation, not the observation that PAFs vary between populations. We therefore harmonised individual-participant data [18] from three nationally representative surveys with contrasting dietary cultures—the USA, Korea and Mexico—and asked three questions. First, is the DII–MetS association comparable across countries once MetS is defined by a single rule? Second, do country-specific PAFs, computed on a common counterfactual scale, differ beyond chance? Third, does the DII’s construct validity—its correlation with C-reactive protein— travel across these settings, and if not, can the observed country ordering survive a formal bias analysis? We did not assume that any cross-national difference exists; the analysis was designed to test whether one can be identified.

## Methods

### Surveys and harmonisation

We used the US National Health and Nutrition Examination Survey (NHANES) 2017–2018 (n=3,391 adults aged 20–80), the Korea National Health and Nutrition Examination Survey (KNHANES) 2015–2021 (n=35,781 adults aged ≥19 across seven cycles) and the Mexican Encuesta Nacional de Salud y Nutrición (ENSANUT) 2018–2019 [28] (n=17,193 adults aged ≥20). All surveys are publicly released and de-identified; the NHANES protocol was approved by the NCHS Research Ethics Review Board and all surveys obtained written informed consent. The harmonised library comprised 56,365 adults (women 32,113).

### Dietary exposure

The energy-adjusted DII (E-DII) was computed from the DII component nutrients available in each survey’s dietary recall (21 components in KNHANES, 32 in NHANES, 20–21 in ENSANUT) using the original algorithm [5]. Because component coverage differs by survey, the index is best interpreted as a survey-specific summary of dietary inflammatory potential rather than as an identical instrument; this is examined directly in the validity analysis below.

### Metabolic syndrome

The primary definition was the harmonised ATP III rule [15,16] requiring at least three of five components: waist circumference above the country-appropriate threshold (US/Mexico >102 cm in men and >88 cm in women; Korea ≥90 cm in men and ≥85 cm in women [12]), triglycerides ≥150 mg/dL, HDL-cholesterol <40 mg/dL in men and <50 mg/dL in women, fasting glucose ≥100 mg/dL or glucose-lowering medication, and blood pressure ≥130/85 mmHg or antihypertensive medication. A four-component definition (waist, triglycerides, HDL-cholesterol, glucose; no BP) was pre-specified as the definition that is computable in all three surveys without BP, and therefore as the test of outcome-definition dependence.

### Recovery of Mexican blood pressure

ENSANUT 2018–2019 BP readings were not included in the pre-harmonised analysis file. We recovered them from the raw anthropometry and hypertension modules: two systolic and two diastolic readings per participant, matched to the harmonised records by household and person keys (16,866/17,193 records matched, 98.1%). Readings coded 222 (the survey’s “not measured” sentinel) were set to missing; implausible values (systolic <60 or >250 mmHg, diastolic <30 or >150 mmHg, or diastolic ≥ systolic) were excluded; the mean of available readings was used. Usable BP was available for 14,637 Mexican participants (85.1%). Antihypertensive medication was not recorded in ENSANUT, so the Mexican BP component is based on measured BP only.

### Statistical analysis

Survey-weighted logistic regression was used throughout, with design variables (strata and primary sampling units) accounting for the complex survey design; KNHANES cycles were modelled with year-indexed strata. Models of MetS included age, sex, body-mass index (BMI) and country, with sex-stratified and country-stratified counterparts. Multiplicative interaction was tested by Wald test. Country-specific PAFs for women were computed on a common counterfactual scale as the survey-weighted fraction of MetS cases attributable to shifting the E-DII distribution to the country-specific lowest-quintile median, using country-specific models adjusted for age and BMI. Confidence intervals and between-country differences were obtained by a survey-design bootstrap (2,000 resamples, resampling primary sampling units within strata) and, as a check, by the delta method. Because the bootstrap difference distribution is bounded and the country samples are independent, we report both the bootstrap intervals for country-specific PAFs and delta-method intervals for pairwise differences.

### Construct validity and quantitative bias analysis

In NHANES and KNHANES (2015–2018, the cycles with hsCRP), we estimated the correlation between the E-DII and log hsCRP by sex, and compared the US and Korean coefficients by Fisher z test. We then asked how much of the observed country difference in the E-DII– MetS odds ratio could be produced by the observed difference in validity. Under the classical measurement-error model, the observed log odds ratio is attenuated by the validity coefficient ρ (the correlation between the measured index and the latent dietary inflammatory construct): log OR_observed ≈ ρ × log OR_true. We computed the disattenuated OR for each country, the value of ρ at which the Korean and US ORs would coincide, and the ratio of disattenuated ORs with its uncertainty. Because the validity coefficients are themselves estimated with uncertainty and because the DII–hsCRP correlation is a proxy for the latent validity, this analysis is presented as an illustration of non-identifiability rather than as a correction to be taken literally.

### Component analysis

To ask whether the composite association was carried by a single MetS component, we modelled each of the five components separately as a function of the E-DII, age and BMI, by country and sex, and compared the component-specific odds ratios. All analyses used R 4.5/4.6 with the survey package [27]. No a priori sample-size calculation was performed; the study is a secondary analysis of large national surveys, and minimum detectable effects are reported in the Supplementary material.

## Results

### Harmonised samples and prevalence

The harmonised library comprised 56,365 adults; after requiring complete data on the five MetS components, 40,521 remained (USA 2,059; Korea 30,077; Mexico 8,385) (Figure 1, Table 1). Weighted MetS prevalence under the common five-component definition was 39.2% in US women, 19.9% in Korean women and 50.2% in Mexican women; the corresponding male prevalences were 41.1%, 25.1% and 36.5%.

**Table 1.**
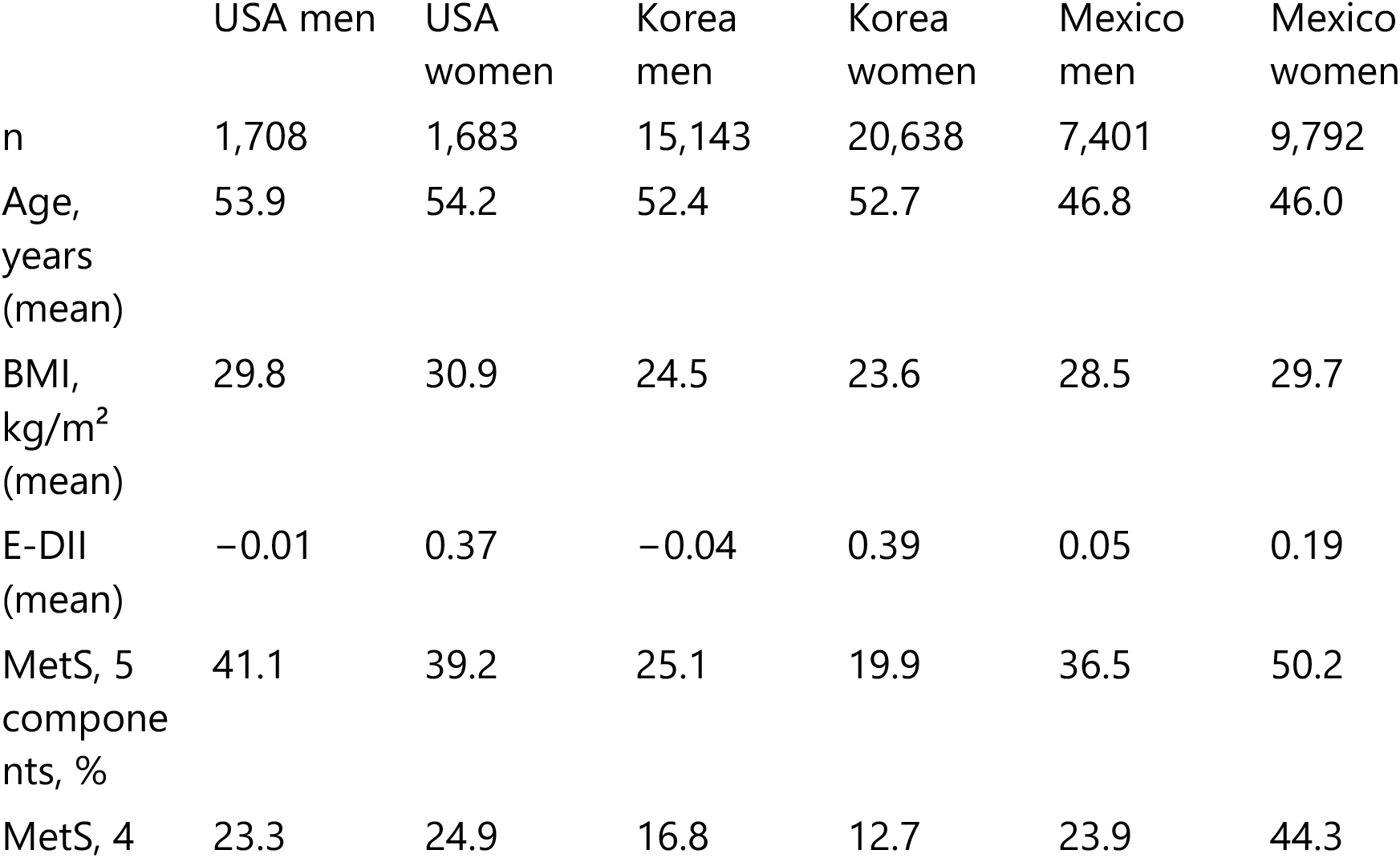
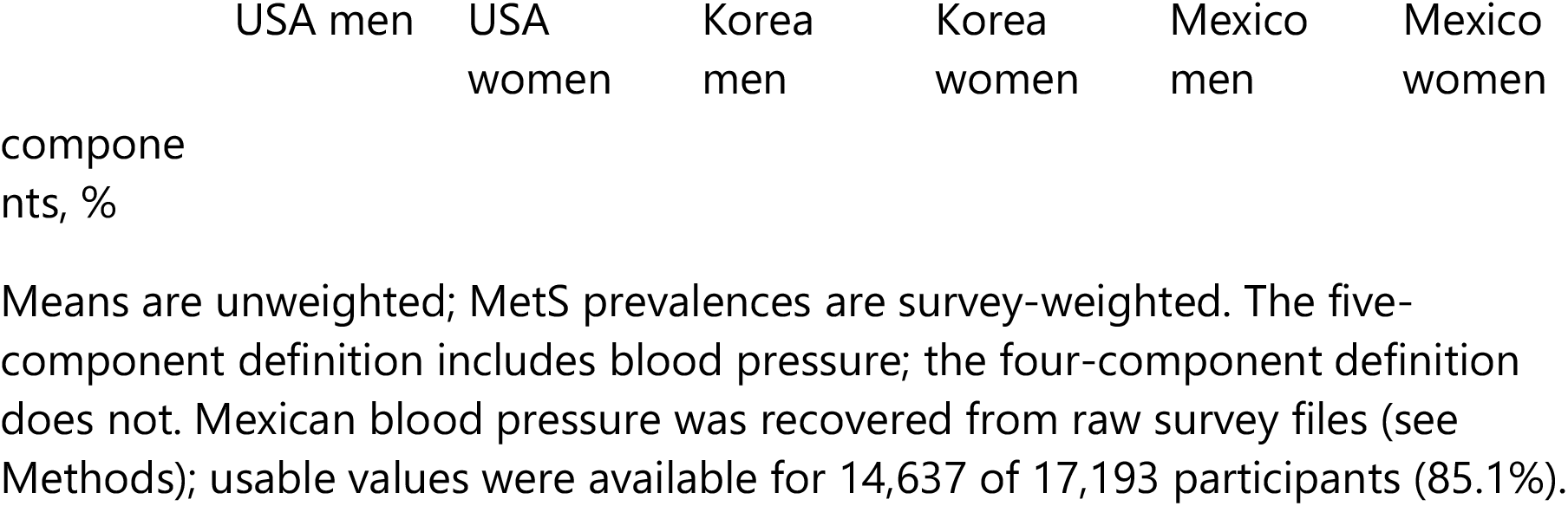
Harmonised sample characteristics, by country and sex.

**Figure 1.** Survey flow and analytic samples for the three national surveys.

### Association under the common definition

Under the five-component definition, point estimates for the E-DII–MetS association were above 1 in women in all three countries, with confidence intervals of 1.23 (95% CI 0.98–1.55; P=0.072) in the USA, 1.078 (1.028–1.132; P=0.002) in Korea and 1.074 (0.992–1.163; P=0.079) in Mexico; the sex interaction was weak (OR 1.083, 0.990–1.184; P=0.080). Male estimates were close to the null (USA 1.021, Korea 1.001, Mexico 1.009) (Table 2). Under the four-component definition without BP, the female association was no longer distinguishable from the null (pooled OR 1.065, 0.982–1.155; P=0.13; country estimates 1.12, 1.036 and 1.067), and the sex interaction was null (P=0.44). The female pooled estimate was 1.065 (0.982–1.155) when BP was excluded from the outcome, and larger when it was included, indicating that the composite association depends on whether BP is part of the outcome.

**Table 2.** Association of the E-DII with MetS, by country, sex and outcome definition (odds ratio per one-unit higher E-DII).

| Definition | Country | Women OR (95% CI) | P | n | Men OR (95% CI) | P | n |
| --- | --- | --- | --- | --- | --- | --- | --- |
| 5 components | USA | 1.23 (0.98–1.55) | 0.072 | 1,053 | 1.021 (0.841–1.239) | 0.821 | 1,006 |
| 5 components | Korea | 1.078 (1.028–1.132) | 0.002 | 17,285 | 1.001 (0.951–1.054) | 0.971 | 12,792 |
| 5 components | Mexico | 1.074 (0.992–1.163) | 0.079 | 4,826 | 1.009 (0.917–1.109) | 0.859 | 3,559 |
| 4 components | USA | 1.12 (0.865–1.449) | 0.359 | 1,068 | 0.971 (0.768–1.227) | 0.787 | 1,023 |
| 4 components | Korea | 1.036 (0.977–1.099) | 0.238 | 17,397 | 1.004 (0.949–1.062) | 0.895 | 12,863 |
| 4 components | Mexico | 1.067 (0.987–1.154) | 0.105 | 5,454 | 1.025 (0.910–1.155) | 0.681 | 4,058 |

Models are survey-weighted logistic regressions adjusted for age and BMI. Sex interaction: five-component OR 1.083 (0.990–1.184), P=0.080; four-component OR 1.047 (0.932–1.177), P=0.436.

### Between-country PAFs

On the common counterfactual scale, the female PAF was 15.0% (95% CI 3.5–25.0) in the USA, 7.6% (3.2–11.9) in Korea and 3.8% (−0.5 to 8.1)† in Mexico. The pairwise differences were 7.4 percentage points (US–Korea; P=0.43), 11.2 (US–Mexico; P=0.23) and 3.8 (Korea–Mexico; P=0.34). No country ordering was statistically distinguishable (Table 3; Figure 2). The point estimates span a factor of about four, but the confidence intervals overlap across their full range, and the Mexican interval includes zero.

**Table 3.** Female population-attributable fractions under the five-component definition and pairwise differences.

| Country | PAF, % (95% CI) | n |
| --- | --- | --- |
| USA | 15.0 (3.5–25.0) | 1,053 |
| Korea | 7.6 (3.2–11.9) | 17,201 |
| Mexico | 3.8 (–0.5 to 8.1)<sup>†</sup> | 4,826 |

| Contrast | Difference, percentage points (95% CI) | P |
| --- | --- | --- |
| USA – Korea | 7.4 (–11.0 to 25.8) | 0.43 |
| USA – Mexico | 11.2 (–7.2 to 29.5) | 0.23 |
| Korea – Mexico | 3.8 (–4.0 to 11.5) | 0.34 |

PAFs are the survey-weighted fraction of cases attributable to shifting the E-DII distribution to the country-specific lowest-quintile median; intervals from the survey-design bootstrap (2,000 resamples); differences by the delta method.
<sup>†</sup>Mexico interval approximate after blood-pressure cleaning. PAF models required a non-missing DII quintile, hence Korea n=17,201 here versus 17,285 in Table 2.

**Figure 2.** Female population-attributable fractions by country (five-component MetS definition), with survey-bootstrap 95% confidence intervals; pairwise differences are given in Table 3.

### Construct validity did not travel

The E-DII–hsCRP correlation among women was 0.137 (95% CI 0.089–0.184; n=1,662) in the USA and 0.040 (0.021–0.058; n=11,215) in Korea (Fisher z P=2.0×10−4); the weighted correlations were 0.134 and 0.018 (Supplementary Table S1). The Korean index therefore tracked circulating inflammation only weakly. Two structural differences are relevant: the KNHANES index was built from 21 components and the NHANES index from 32, and the Korean diet differs in the foods that dominate the index.

### Bias analysis

The ratio of validity coefficients (Korea/USA) was 0.294 (95% CI 0.164–0.528). The value of the Korean validity coefficient at which the disattenuated Korean and US odds ratios would coincide was 0.055, which lies inside the confidence interval of the observed ratio; the disattenuated OR ratio (Korea/USA) was 1.46 at the observed Korean coefficient (0.040) and crossed unity at a coefficient of 0.050, so the observed ordering (Korea below the USA) was not preserved. The ratio of validity coefficients (0.29, 95% CI 0.16–0.53) straddles the reversal threshold (0.37), so the direction of the country difference was not identified. The analysis therefore does not identify a country ordering: depending on the validity assumed, Korea can be below, equal to, or above the USA (Figure 3).

**Figure 3.** Sensitivity of the disattenuated Korea/USA odds-ratio ratio to the assumed Korean validity coefficient. The horizontal line marks equality; the vertical line marks the observed Korean coefficient (0.040) and the shaded region the confidence interval of the validity ratio.

### Component analysis

The association was not carried by BP: the female BP-component odds ratio was 1.195 (0.936–1.525) in the USA, 1.029 (0.992–1.068) in Korea and 1.075 (0.979–1.180) in Mexico, all non-significant; adjusting the five-component MetS model for BP attenuated the female association by only 12.4% (1.105 to 1.091; P=0.011). The component that was consistently associated in all three countries was low HDL-cholesterol (USA 1.265, 1.076–1.487; Korea 1.055, 1.016–1.096; Mexico 1.076, 1.006–1.151; pooled 1.119, 1.063–1.178). The composite association therefore reflects a component whose weight in the composite differs across settings, not a single dominant pathway.

## Discussion

In three nationally representative surveys, harmonised at the individual level, we found that the cross-national ordering of diet-attributable MetS burden was not identified. The point estimates of female PAFs differed several-fold, but no pairwise difference was statistically distinguishable; the association itself was present or absent depending on whether BP was included in the MetS definition; and the index’s correlation with C-reactive protein differed more than threefold between the USA and Korea, enough that a formal bias analysis could reverse the country ordering. These findings do not support the transport of diet-attributable burden estimates across dietary cultures. They suggest instead that cross-national comparisons must demonstrate, rather than assume, that the index and the outcome mean the same thing in each population.

The result that the association is definition-dependent is not a technicality. MetS is a composite whose components are themselves outcomes of different aetiologies, and the DII may be more strongly related to one component (here, HDL-cholesterol) than to others. When the composite is assembled differently— for example, without BP in a survey that did not measure it—the association changes. This is the mechanism by which the same data can yield 1.105 or 1.065, and it explains why the published DII–MetS literature reports estimates that move with the definition used [8]. The finding also argues against reading a composite association as evidence of a single biological pathway. That the association was larger in women is consistent with sex-hormone modulation of inflammatory pathways [20–23] and with the stronger inflammatory prediction of incident diabetes in women [24–26].

The validity finding is the more consequential one for cross-national work. The DII was developed and validated in Western populations, where its correlation with CRP is modest (r≈0.2–0.3) [6,19]. In our data the correlation was 0.137 in the USA but 0.040 in Korea, and the weighted Korean estimate was 0.018. An index that does not track inflammation in a population cannot be used to compare that population’s inflammatory-diet burden with another’s. The quantitative bias analysis makes the consequence explicit: the observed country ordering is compatible with a true ordering in either direction, and the disattenuated PAFs diverge rather than converge because baseline MetS prevalence also differs. This is not a problem that more precise estimation can fix; it is a problem of construct validity, and it requires local calibration of the index before PAFs are compared.

Our findings reframe, rather than contradict, the single-country literature. The positive association reported in Korean men [13] and the high hazard ratio reported in a Mexican cohort [14] are single-population estimates that we cannot reproduce or refute with cross-sectional data; our point is that their magnitudes cannot be compared with one another, or with US estimates, without evidence that the index and the outcome are commensurable. The same caution applies to the meta-analytic literature, which pools study-level estimates across countries and definitions [7–9]. Pooling assumes commensurability; our data show that the assumption can fail in both the exposure and the outcome.

The study has several limitations. The design is cross-sectional, so the estimates describe associations, not effects; PAFs assume causality and are presented as hypothesis-generating quantities [17]. Mexican BP was recovered from raw survey files and cleaned of sentinel codes, but antihypertensive medication was not recorded, so the Mexican BP component is measured differently from the US and Korean components, in which medication data were partly available. The US sample contributed only 1,053 women with complete five-component data, which is why the US estimates are imprecise. The DII was built from different numbers of components in each survey, and the hsCRP correlation is a proxy for the latent validity of the index rather than a direct measurement-invariance test. Finally, the bias analysis uses a classical measurement-error model whose disattenuated estimates are unstable when validity is low; we use it to demonstrate non-identifiability, not to produce corrected PAFs.

## Conclusion

In harmonised individual-participant data from three dietary cultures, the cross-national ordering of diet-attributable metabolic burden was not identified. The apparent differences were bounded by the definition of the outcome and by the validity of the dietary index, and neither bound is removable by analysis alone. Cross-national comparisons of diet-attributable burden should report the validity of the index and the comparability of the outcome in each population, and should treat country rankings as unidentified until both are demonstrated.

## Supporting information

Supplementary Table S1 and MDE

## Data Availability

Supplementary results (CSV format), analysis scripts (R version 4.6.1, Python version 3.13), and aggregated data are provided as Supplementary Material accompanying this manuscript. GWAS summary statistics: CRP GWAS (ebi-a-GCST90029070; Said et al., 2022) and MetS GWAS (ebi-a-GCST90444487-9; Park et al., 2024) are available at https://gwas.mrcieu.ac.uk and https://www.ebi.ac.uk/gwas/. LD clumping used the 1000 Genomes Project Phase 3 European reference panel (EUR, n=503). DII was calculated using the global reference database of Shivappa et al. (2014, Public Health Nutrition).

https://www.ebi.ac.uk/gwas/

## Declarations

### Ethical approval

All three surveys were approved by their respective institutional review boards and obtained written informed consent; the present secondary analysis of de-identified, publicly released data required no further ethical approval.

### Data availability

The national surveys are publicly available (NHANES, https://www.cdc.gov/nchs/nhanes/; KNHANES, https://knhanes.kdca.go.kr/; ENSANUT, https://ensanut.inspq.mx/). The harmonised analysis dataset and analysis code are available from the corresponding author and will be deposited in a public repository on acceptance.

### Author contributions

All authors contributed to the conception and design of the study, the acquisition and analysis of data, and the drafting and critical revision of the manuscript; all authors approved the final version.

