## Supplementary Table S1 and MDE for "Outcome definition and index validity in cross-national comparisons of diet-attributable metabolic burden"

### Supplementary material

#### Supplementary Table S1. E-DII construct validity and quantitative bias analysis (women).

| | USA | Korea |
|---|---|---|
| E-DII–log hsCRP r (95% CI) | 0.137 (0.089–0.184) | 0.040 (0.021–0.058) |
| n | 1,662 | 11,215 |
| Weighted r | 0.134 | 0.018 |
| Validity ratio Korea/USA (95% CI) | — | 0.29 (0.16–0.53) |
| Corrected OR | 4.52 | 6.62 |
| Corrected OR ratio Korea/USA | — | 1.46 |
| ρ at which the ratio crosses unity | — | 0.050 |

Correlations use log-transformed hsCRP. Corrected estimates apply the classical measurement-error model log OR_observed ≈ ρ × log OR_true; they are presented to illustrate non-identifiability, not as corrected population estimates.

#### Supplementary Methods. Minimum detectable effect

The study was a secondary analysis of national surveys, and the sample size was determined by the surveys rather than by an a priori calculation. With the available female analytic samples and observed MetS prevalence, the minimum detectable odds ratio per one-unit higher E-DII at α=0.05 (two-sided) and 80% power was approximately 1.15 in the US sample (n=1,053), 1.03 in Korea (n=17,285) and 1.06 in Mexico (n=4,826); the corresponding detectable effects are wider for the four-component definition. These values are provided to aid interpretation of the confidence intervals, not as a justification of sample size.
